# Pandemic-era sports league operations as a new paradigm for local epidemic resilience

**DOI:** 10.1101/2025.10.24.25338752

**Authors:** Yun Tao, C. Brandon Ogbunugafor, Nita Bharti

## Abstract

Strategic, coordinated, and rapid responses are essential when pathogens emerge, yet these responses are often mishandled during epidemics due to myriad factors, including entrenched socioeconomic and political constraints. Consequently, local communities are often left unprotected. Early in the 2020 COVID-19 pandemic, the National Basketball Association (NBA) implemented a variation of standard public health measures, popularly termed the “bubble”, that kept the league transmission-free for the 2020 playoffs. The NBA “bubble” was a sophisticated system of risk stratification predicated on a strategic design of spatial, movement, and contact heterogeneities. Five years later, we examine its fundamental features and explore its potential application to future epidemics. We formalize the bubble’s mechanisms in the context of existing epidemiological theory, generate forecasts of management outcomes, and propose a theoretical framework that redirects epidemic modeling away from its traditional top-down control philosophy toward targeted protection and local resilience. The new framework can ultimately be used to support the design and local governance of community protections, particularly when centralized epidemic responses prove inadequate.

## Introduction

Recent epidemics have exposed the limits of traditional, population-wide control strategies. Policy missteps, patchwork implementation, and uneven adherence to public health guidance allowed outbreaks to gain momentum [Bambra, 2022,Bergquist et al., 2020,Wilder-Smith and Qureshi, 2020]. Once transmission surges, two classes of subpopulations can arise that are especially in need of heightened protection. The first comprises clinically vulnerable groups, i.e., those whose age, comorbidities, or immunological status confer a disproportionate risk of severe illness [Doherty et al., 2016,Russell et al., 2023]. The second consists of individuals who sustain critical societal functions during a crisis: healthcare workers, educators, food-supply personnel, and other essential staff whose professional exposure amplifies their risk of infection [Mutambudzi et al., 2021,Scarpino et al., 2016,Selvaraj et al., 2018]. Safeguarding such groups is crucial to public health stewardship and the continuity of operational norms, yet it falls outside the scope of current epidemiological theory, which mostly focuses on reducing epidemic impacts at a large population scale.

In light of such epidemic realities, it is essential to broaden traditional disease modeling paradigms and evaluate small, subpopulation-scale strategies that deliver targeted protection under widespread transmission. In principle, this requires explicitly modeling field-tested instances of these strategies, documented in empirical observations and historical case studies. We are primarily motivated by the following questions: (i) What control mechanisms may be necessary to achieve effective protection of target groups? (ii) Can we predict the strength and duration of targeted protection that a given strategy confers? (iii) To what extent can the resulting insights generalize into a novel epidemic management framework?

Analogous problems involving the protection of target groups have long existed in ecological and general conservation research. Studies of metapopulation ecology suggest that when the broader environment becomes too fragmented and unfavorable, isolating high-value inhabitants can delay large-scale extinction [Johst et al., 2002,Ovaskainen et al., 2002,Tao et al., 2024]. In movement ecology, models increasingly explore the use of physical barriers and corridors to steer at-risk species safely across complex landscapes [Bastille-Rousseau and Wittemyer, 2021,Tao et al., 2025,Unnithan Kumar and Cushman, 2022]. In disease ecology, there has been an emphasis on the value of population buffering, whereby incompetent host species dilute transmission to those that are vulnerable (i.e., dilution effects; see Keesing and Ostfeld [2021], Rohr et al. [2020]). Across these examples, a common principle emerges: targeted protection requires stratifying risk, achievable by engineering heterogeneity in space, movement, and contact to shield the components that matter most.

The COVID-19 pandemic provided real-world confirmation of this notion. Hospitals enforced strict entry screening to protect high-acuity spaces (e.g., Jaswaney et al. [2022]); age-based vaccination schemes were proposed for keeping essential services running while limiting wider spread (e.g., Buckner et al. [2021]); and schools adopted staggered schedules to break transmission chains (e.g., Fong et al. [2020]). In each instance, strategic risk stratification functioned as firewalls around critical subpopulations even as community transmission continued. These control measures suggest that stratification “by design” can complement or, in many contexts, outperform uniform, top-down measures. Integrating this idea into epidemic forecasts could provide an alternative modeling framework that shifts the goal from suppressing transmission everywhere to targeting defensive effort somewhere, namely, on critical subpopulations whose infections would exact the greatest societal cost.

A compelling template for such a framework was exemplified by the National Basketball Association (NBA)’s COVID-19 response. In July 2020, after a mid-season suspension of play prompted by player infections, the NBA resumed its season inside a biosecure “bubble” at Walt Disney World in Orlando, Florida [Golliver, 2021]. All participating players, coaches, staffers, and accredited media members were confined on-site until the end of their stay. A multilayered health and safety protocol was implemented, which included daily testing, tiered grouping of personnel with corresponding housing and access restrictions, tightly monitored, limited movement between tiers, stringent contact regulations, and swift, severe penalties (e.g., expulsion from the bubble) for rule violations [Golliver, 2021]. The result was striking: Zero COVID-19 cases were detected after arrival quarantine in the players or the team staff throughout the event [Mack et al., 2023]. The NBA bubble soon became the exemplar for outbreak management in professional sports; similar protocols were successfully adopted by other leagues, including the Women’s National Basketball Association (WNBA) and the National Hockey League (NHL) [Harshaw, n.d.]. Above all, the NBA bubble stands as a rare modern example in which commercial interests and public health practice overlapped, marking a brief but notable alliance between corporate stakeholders and the scientific community. Five years on, we revisit this landmark social experiment as a conceptual foundation for a new class of epidemic models centered on risk stratification. We first identify key mechanisms of bubble defense, formalizing and integrating them into a theoretical framework. Next, we test this framework in the form of a spatially explicit agent-based model. By modeling how bubbles operate as a general defense strategy, we demonstrate its potential application in everyday settings (e.g., classrooms, workplaces, hospitals) where objectives, resources, and oversight are highly variable.

### Bubble philosophy

A chief innovation of the NBA bubble was its hierarchical application of classical concepts from epidemiology and ecology: members were grouped by management priority, and each level of priority (tier) was paired with a strategic rule set that determined how the individuals were arranged in space, where they could move, and with whom they could interact. In effect, the bubble managed its internal population heterogeneously, with the highest-priority members receiving the strongest protection. This approach echoes the growing number of disease models that identify heterogeneity as a first-order determinant of epidemic dynamics. For instance, non-uniform density distribution and assortative social mixing patterns were found to alter forecasts [Bansal et al., 2007,Manna et al., 2024], and social-economic inequality presents major challenges to outbreak control efforts [Marmot, 2005,Surasinghe et al., 2024,Zelner et al., 2022]. In most studies, heterogeneity is treated as a pre-existing feature of the system that models should take into account, or sometimes an end-state for guiding scenario planning, e.g., setting target reproductive numbers for distinct groups. In contrast, the NBA bubble leverages heterogeneity as a core design element in its management strategy, creating it on demand to provide immediate, targeted protection of mission-critical individuals.

In addition, within the NBA bubble’s structural complexity is a standard disease control playbook, specifically, the use of non-pharmaceutical interventions (NPIs) to limit transmission events, a category of strategies long considered effective in controlling epidemics (e.g., SARS, H1N1 influenza, Ebola) [Bell et al., 2006,Cowling et al., 2020,Merler et al., 2015]. NPIs typically act by reducing contacts between infected (I) and susceptible (S) individuals. An extreme example is the stay-at-home (SAH) order, which reduces interpersonal interactions overall. SAH is used as a blanket strategy when infected individuals cannot be easily identified. In a “coinflip” analogy, where each instance of heads represents an infectious (S-I) contact, SAH minimizes the number of flips (i.e., attempted contacts). Historically, this broad-scale strategy has often amounted to a blunt, heavy-handed response to emerging outbreaks; however, uptake can vary and compliance can wane under prolonged or onerous restrictions [Garnier et al., 2021,Joshi and Musalem, 2021]. The COVID-19 pandemic highlighted an additional challenge: specific cohorts, such as healthcare providers, high-risk patients, and essential workers, need to maintain frequent contacts during a crisis [Nelson et al., 2022,Wilson-Aggarwal et al., 2022]. Thus, scenario planning for NPIs, SAH included, faces intrinsic scale limitations: the population that can be realistically covered and the duration over which it can be sustained. In contrast to SAH, bubbles are intended to provide localized interventions within an uncontrollable or unpredictable general population. By applying NPI policies to only select individuals and contact types, they form semi-closed subpopulations for which the outcome of every “coin-flip” – not the number of flips – is manipulated to favor non-transmission, marking a philosophical shift in epidemic management.

At first glance, a bubble-based strategy seems paradoxical: it protects individuals by redistributing them in a densely interconnected network. If infection were introduced, the number of secondary infections, as measured by the basic reproductive number R0, could soar, contradicting the standard management goal of keeping R0 below unity. However, we note that the principle behind the bubble is not to sever contacts indiscriminately, but to shield its members from external threats. By tightly controlling all potential points of pathogen entry and rapidly detecting infections within, interactions that would be risky outside the bubble can proceed relatively safely inside it. As long as every member is infection-free, the volume of internal contacts matters far less than it does under SAH.

Since only a subset of the wider population is given this targeted protection, the bubble approach is non-egalitarian in nature. Eligibility rests on context-specific objectives and priorities, as determined by functional necessity (e.g., participants in the NBA’s return-to-play, students attending classes) or increased protection for vulnerable groups (e.g., immunocompromised individuals, specific age classes). Bubble formation entails many layers of demographic and ethical considerations.

Beyond safeguarding high-risk or mission-critical individuals, bubbles confer several practical advantages. When applied in public settings, such as a multi-household network, they can restore social interactions that prolonged SAH would preclude, buffering the mental-health toll of isolation, especially in children [Meade, 2021]. From an operational standpoint, bubbles are highly flexible. They can be designed around timelines and resource constraints, including the goodwill of the participants. When the end date is fixed and transparent (e.g., the final day of an NBA season), intensive measures such as bio-surveillance and contact tracing can be carried out with managed long-term expectations for cost and compliance. Conversely, bubbles designed to operate indefinitely (for example, remaining in place until disease prevalence falls below a predefined threshold) may be sustained by limiting their membership and periodically easing costly oversight in exchange for continued cooperation, with clear tradeoffs for effectiveness. Within a bubble, every detected infection should prompt immediate expulsion to prevent uncontrolled spread. A bubble contracts over time unless the removed members are replaced, a process termed “relational exchange” [Scarpino et al., 2016]. The likelihood of a breach that results in pathogen importation scales with the external force of infection, particularly in neighboring communities. General population-scale measures that lower community prevalence (e.g., SAH) can therefore promote a bubble’s success by reducing import pressure. During COVID-19, several countries adopted a similarly mixed strategy, coupling small bubble-like groups (e.g., pods) with broad lockdowns [Clayton et al., 2020,Rankin, 2020].

The bubble concept introduces unique management features that can extend current epidemiological theory and enhance our epidemic preparedness. Its mechanisms may be organized around three key features: 1) semi-insulation, 2) boundary control, and 3) contact assortativity. To date, no dedicated model of bubble strategies has been developed, yet the necessary building blocks can be found scattered across disparate strands of the literature. In this paper, we assemble these mechanistic fragments into conceptual and theoretical frameworks, laying the groundwork for future research on bubble-based control and, more broadly, providing a new, scalable tool for scenario planning in an increasingly complex world.

## Results

### Conceptual overview

Our first contribution to the subject is conceptual: in the simplest form, a bubble is a physical or social barrier that fences a subpopulation, affording it a certain degree of insulation from the surrounding epidemic landscape while still admitting some logistical traffic (Fig 1a). Within a compartmental model framework, entry into a bubble resembles quarantine: both require initial screening and temporary withdrawal from the general population. The purpose, however, is inverted: quarantine protects the community (out-group) from the entrants (in-group), while bubbling protects the entrants from the community. Moreover, quarantined individuals are typically already exposed or infectious, while bubble members are ideally still susceptible or immune. Bubble members’ interactions with all individuals outside the bubble, including uninfected individuals, are restricted overall and quantified by porosity.

**Fig. 1.**
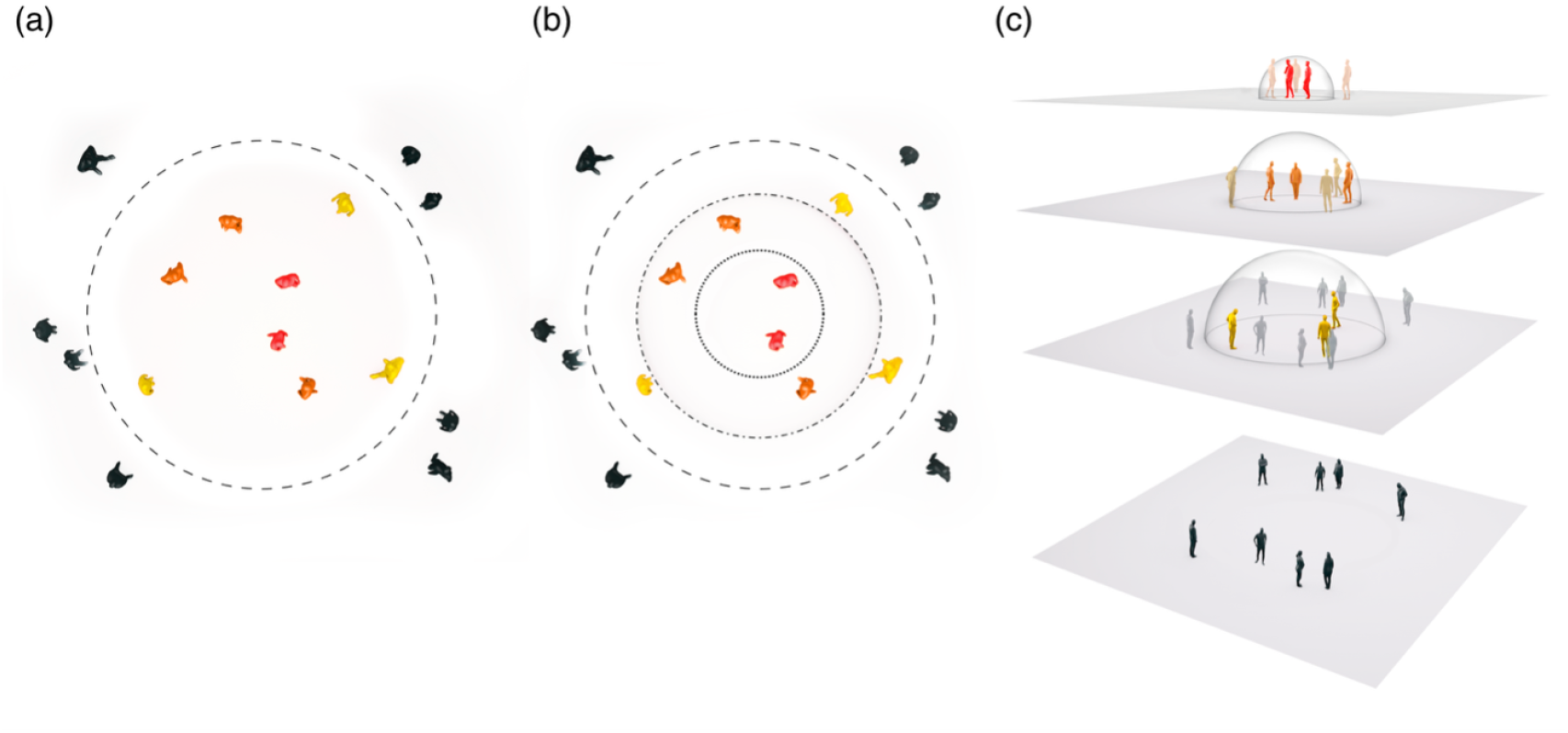
Conceptual schematics of bubble architectures: (a) a single-tier bubble, (b) a multi-tier bubble, and (c) a multi-tier bubble visualized in three-dimensional space with overlapping planes of interaction. Individuals differ in their priority for protection, ranging from lowest (black) to highest (red). In (a) and (b), adjacent tiers are separated by bubble boundaries that vary in permeability: mildly porous (dashed line), moderately porous (dash-dotted line), and highly porous (dotted line). For visual clarity, only individuals in the focal tiers and their nearest-outer (lower) tiers are shown in (c).

A defining feature of bubble defense is the partition of infection risk across population strata. In the model above, stratification is achieved implicitly by a single, semi-porous boundary; a perimeter with controlled access points that separates members from non-members. Porosity can be further reduced by nesting the bubble, that is, dividing it into hierarchical tiers that provide finer separation of higher-priority individuals from outer tiers where contacts occur more freely. In a deterministic model, these tiers can be represented as concentric shells, each defined by its geometric distance from the background environment: the general population occupies tier 0, while the most protected group resides in tier n (Fig 1b). In settings where bubbles arise from modified social behaviors rather than physical barriers, the structural hierarchy may be better pictured as a three-dimensional network of interaction planes (Fig 1c). This view avoids the impression of rigid internal boundaries and makes clear that same-tier contacts are not spatially constrained.

Multiple bubbles can also exist on a single landscape, forming a mosaic of variably nested, regionally overlapping population “islands”. As the number of bubbles grows, community patchiness may increase, analogous to the effect of habitat fragmentation. Recent advances in metapopulation theory, when interpreted in the context of epidemiology, suggests that substituting one large bubble with several smaller ones can drastically alter longterm pathogen persistence, disease prevalence in the total population, and spatial synchrony of local outbreaks [Tao et al., 2024].

Early case detection and boundary security are necessary for curbing onward transmission, but when detection fails and boundaries are “leaky”, contact assortativity can provide as an additional guardrail by increasing the predominance of preferential mixing, i.e., concentrating contacts among individuals from the same tier of origin, while limiting proportionate mixing, in which contact rates depend solely on local densities. Both preferential and proportionate mixing can be parameterized in an epidemic model and controlled in response to management risk tolerance. For example, on suspicion of new cases, the ratio of preferential to proportionate mixing can be temporarily increased to contain potential spread. Conversely, it can be reduced during periods when more unrestricted or cross-tier contacts are required (e.g., sporting tournaments, school hours).

Until the external epidemic is brought under control, even a well-insulated, functionally stratified, and highly assortative bubble will likely need to be supported by strong oversight, role redundancy, and rapid response. Strong oversight entails continuous surveillance of infection status, contact tracing, and enforcement. Because detection is never perfect, some violations of bubble rules inevitably go unobserved or unpenalized. Minimizing the frequency of such lapses helps prevent cryptic chains of transmission from forming inside a bubble. Role redundancy is essential for balancing safety against functions. If every infected or even suspected case is expelled immediately, transmission risk decreases but essential tasks may stall. While professional sports leagues address this problem with roster depth, many social bubbles lack such built-in mechanisms for role replacement. To stay viable, they must intentionally include redundancies for each key role so that essential activities can continue when some members are removed, an idea known as the “bus factor” in software project development [Cosentino et al., 2015]. Allowing the bubble to contain, or be reloaded with, not only susceptibles but also immune or recovered individuals could further enhance its long-term viability. Rapid response anchors every line of bubble defense. Once a pathogen breaches the bubble perimeter, any delays stemming from diagnostic or reporting lags, logistical bottlenecks, or patchwork management can markedly worsen outbreak severity (see Tao et al. [2021]). A clear understanding of the bubble’s operational infrastructure, and of its hidden fracture points, allows managers to allocate resources strategically and mitigate sources of delays.

The feasibility of any bubble design and its supporting measures ultimately hinges on resource availability. Unlike major U.S. sports leagues, most community bubbles cannot fund expert modelling, extensive screening, and professional intervention protocols. Instead, they must rely on self-planning and coordination, voluntary participation and honest reporting, and other “grassroot” measures, all of which are typically less reliable. During the early years of COVID-19, households were encouraged to create self-regulated social bubbles. Most of these makeshift arrangements lacked resources and were uninformed on proper bubble practices. In economically disadvantaged communities, pandemic-related labor pressures also made adherence difficult [Chang et al., 2021,Jay et al., 2020]. Under these conditions, simple, low-cost bubbles can be designed by limiting population size, the number of tiers, and oversight demands. Exploring the economic dimensions of the most critical protective elements could thus help identify strategies that could be implemented and self-organized even under severe resource constraints.

### Agent-based model: a pilot analysis

Our second contribution is theoretical: as a simple demonstration of the bubble framework, we developed a spatially explicit agent-based simulation model incorporating the mechanisms described above. The model represents a bubble composed of three concentrically nested tiers (T1, T2, and T3), embedded within a general population (GP) during an emerging outbreak (Fig 2). All members are initially infection-free, and individuals in higher (more inner) tiers are more spatially proximate thus interact more frequently. The management goal is to prevent pathogen introduction or, failing that, to minimize disease prevalence in tiers close to the bubble center.

**Fig. 2.**
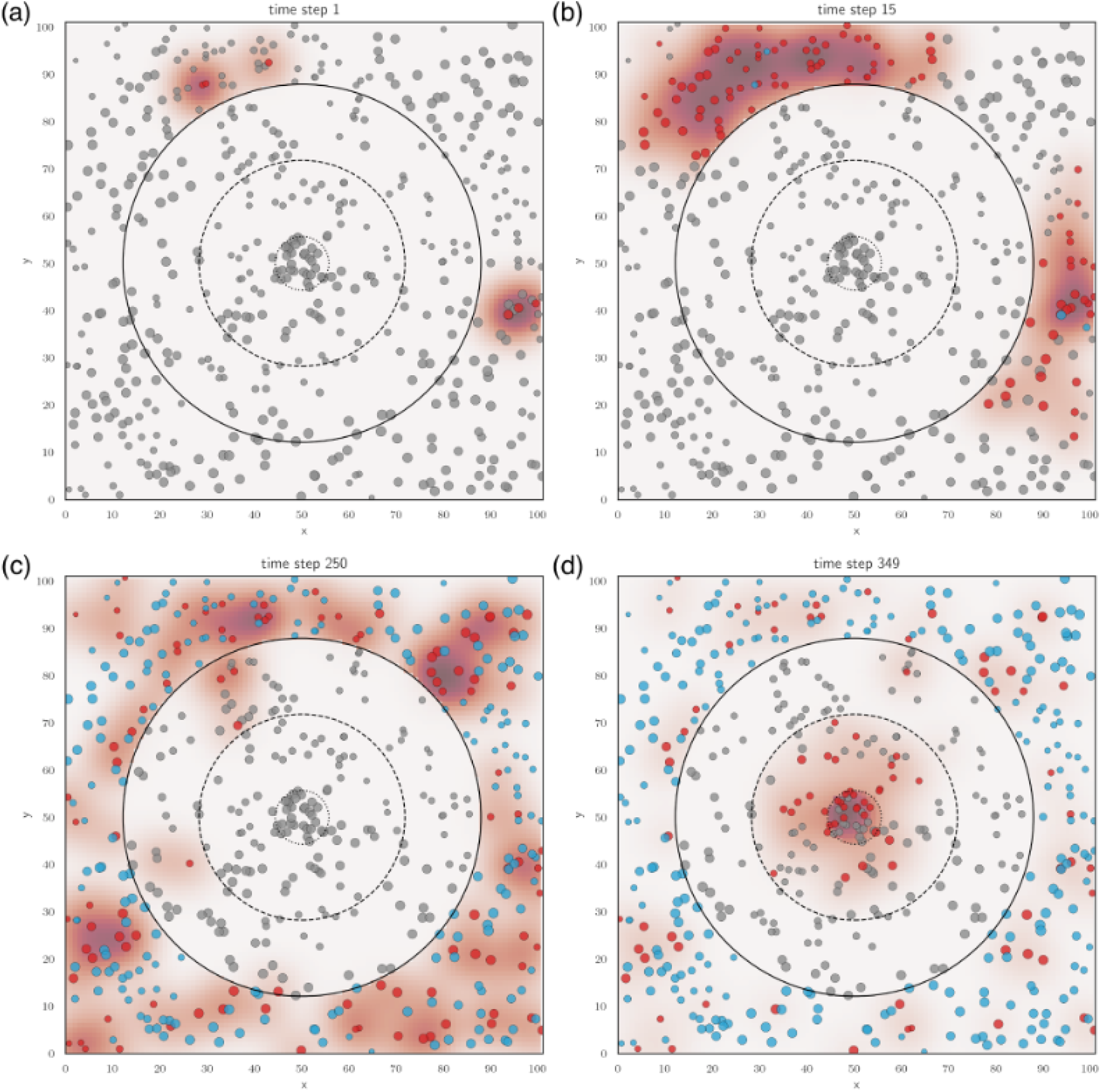
Agent-based simulation of a three-tier bubble defense on a 101 × 101 lattice. The population is partitioned into the general population GP (*n* = 300) and three nested tiers: T1 (*n* = 110), T2 (*n* = 70), and T3 (*n* = 30). (a-d) Snapshots of an outbreak seeded in GP. Individuals are indicated by circles and coloured by disease state (grey: susceptible; red: infected; blue: recovered). Background color shows the local forces of infection after each cycle of transmission (before recovery and relational exchange). Contacts follow a bivariate-Gaussian spatial kernel (width *α* = 3). Individual rates of infection *β*, depicted by circle sizes, are randomly drawn from a spatially correlated field ranging from 0.5 to 25. In GP, recovery rate *γ* = 0.005. In T1-T3, relational exchange rate *ζ* = 0.6. Boundary porosities *ω*_0,1_ = *ω*_1,0_ = 0.1, *ω*_1,2_ = *ω*_2,1_ = 0.2, and *ω*_2,3_ = *ω*_3,2_ = 0.5. Mixing is entirely proportionate (*ϵ* = 0).

We assume all individuals are mobile, and transmission occurs when an infected individual encounters a susceptible individual at any location. To capture this, we auto-convolve a Gaussian interaction kernel to represent the meeting probability of two independently moving individuals. The resultant kernel is then convolved with the spatial distribution of infected individuals in each tier, modulated by boundary porosity, to obtain tier-specific contributions to local forces of infection. The contact structure comprising both proportionate and preferential mixing operates identically to the well-mixed, non-spatial formulation in Eqs. (2)-(4), but is evaluated pointwise over the simulated landscape. More specifically, the proportionate mixing component is weighted by contact opportunity, which we quantified via the kernel-smoothed density of infectious individuals from each source tier at the susceptible’s location, rather than by the mean density of that tier. This preserves the conceptual structure of the analytical equations while allowing heterogeneous population density to influence transmission risk.

Members are behaviorally heterogeneous in their adherence to bubble rules. We model this by letting transmissibility vary spatially, so that individuals in nearby locations experience similar levels of risk. Infected members are handled through relational exchange; new members carry random adherence values, which are then spatially averaged with those of their neighbors.

Our simulation results revealed several notable patterns. First, once the bubble is breached, infections surface in the outermost tier and cascade inward, often with brief delays between tiers (Fig 3). Second, since interactions intensify toward the center, prevalence rises with each tier level (Fig 3). Third, bubble effectiveness can vary drastically based on design parameters. A poorly implemented bubble – characterized by slow response, highly permeable boundaries, and indiscriminate mixing – offers little protection: its members become rapidly infected as community transmission surges; early on, the innermost tier (T3) may even suffer higher prevalence than the general population, rendering the bubble counterproductive (Fig 3a). However, modest strategic adjustments can yield meaningful improvement. Accelerating the response alone significantly reduces within-bubble prevalence (Fig 3b). Imposing tighter boundaries between tiers extends the bubble’s lifespan (Fig 3c). Finally, incorporating preferential mixing on top of these interventions achieves long-term bubble security (Fig 3d).

**Fig. 3.**
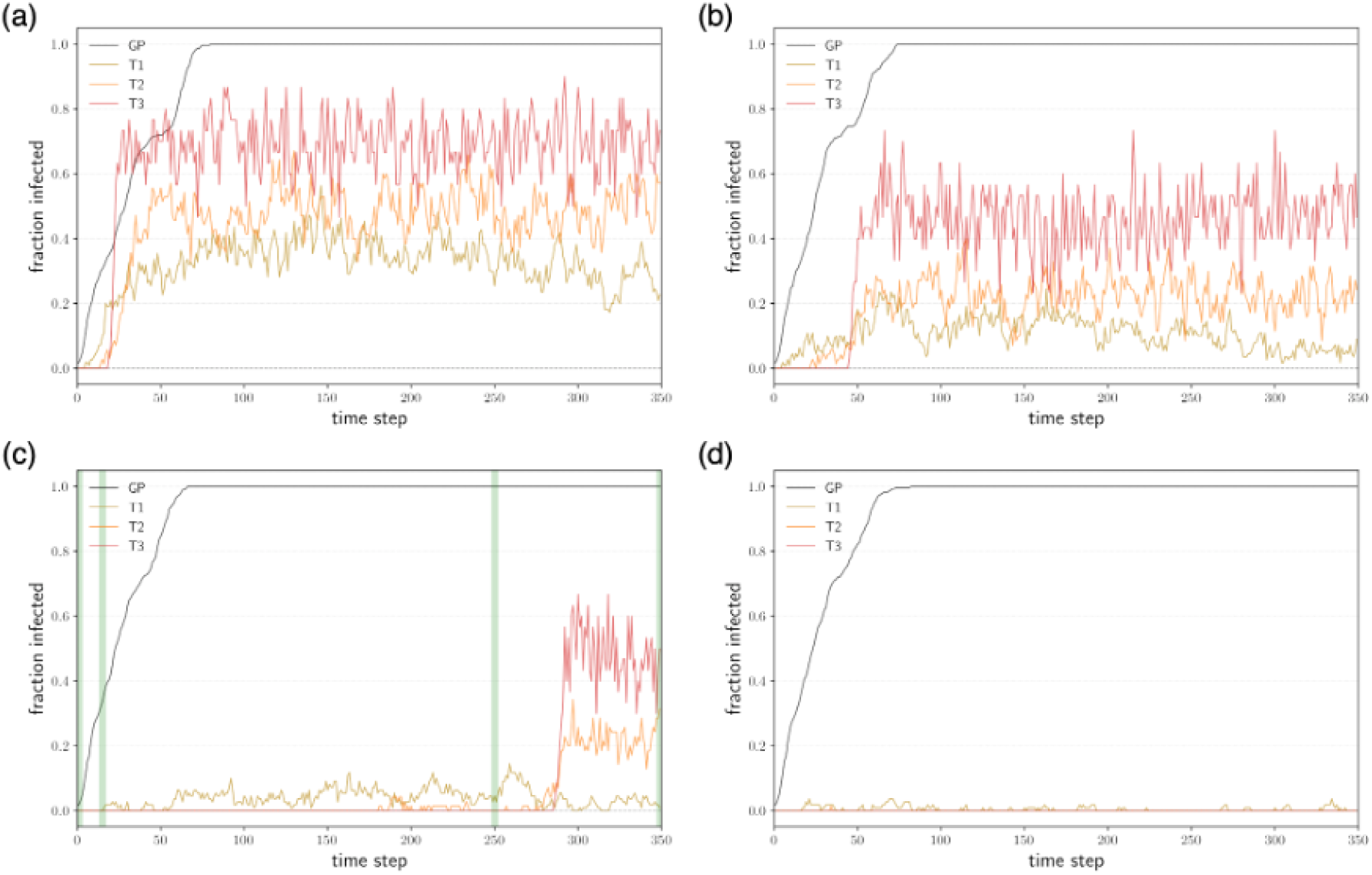
Tier-specific prevalence under increasingly protective regimes. Bubble architecture, model processes, and model parameters are identical to those in Fig. 3 unless noted otherwise. (a) Weak protection: relational exchange rate *ζ* = 0.3; uniform boundary porosity *ω*_*i, j*_ = 0.6; no preferential mixing (*ϵ* = 0). (b) Moderately weak protection: as in (a) but with *ζ* = 0.6. (c) Moderately strong protection: as in (b) but with heterogeneous boundary porosities, identical to the bubble configuration in Fig. 2. Green vertical bars correspond to the timestamps shown in Fig. 2a-d. (d) Strong protection: as in (c) but with preferential-contact matrix 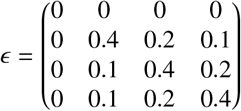, rows/columns ordered GP → T1 → T2 → T3.

If infected members are removed and not replaced (e.g., re-entry is denied after recovery), a bubble exposed to sustained external transmission will eventually empty via stochastic pathogen importations, akin to the inevitability of local extinctions in metapopulation theory [Ovaskainen and Hanski, 2004,Tao et al., 2024]. Membership declines more slowly in a well-defended bubble, where boundary porosity is low and assortativity is high, compared to a bubble with weaker protections. The bubble’s internal population dynamics therefore correlate with its performance. We can measure this population decline by modeling the time-to-implosion *T*_*imp*_, the time at which the fraction of members in any tier drops below an operational threshold, such as the minimal roster size per team required to continue a sporting tournament or a functional community size (Fig 4). We expect a negative relationship between *T*_*imp*_ and disease persistence in the general population: poor control of widespread outbreaks shortens *T*_*imp*_, undermining long-term bubble viability.

**Fig. 4.**
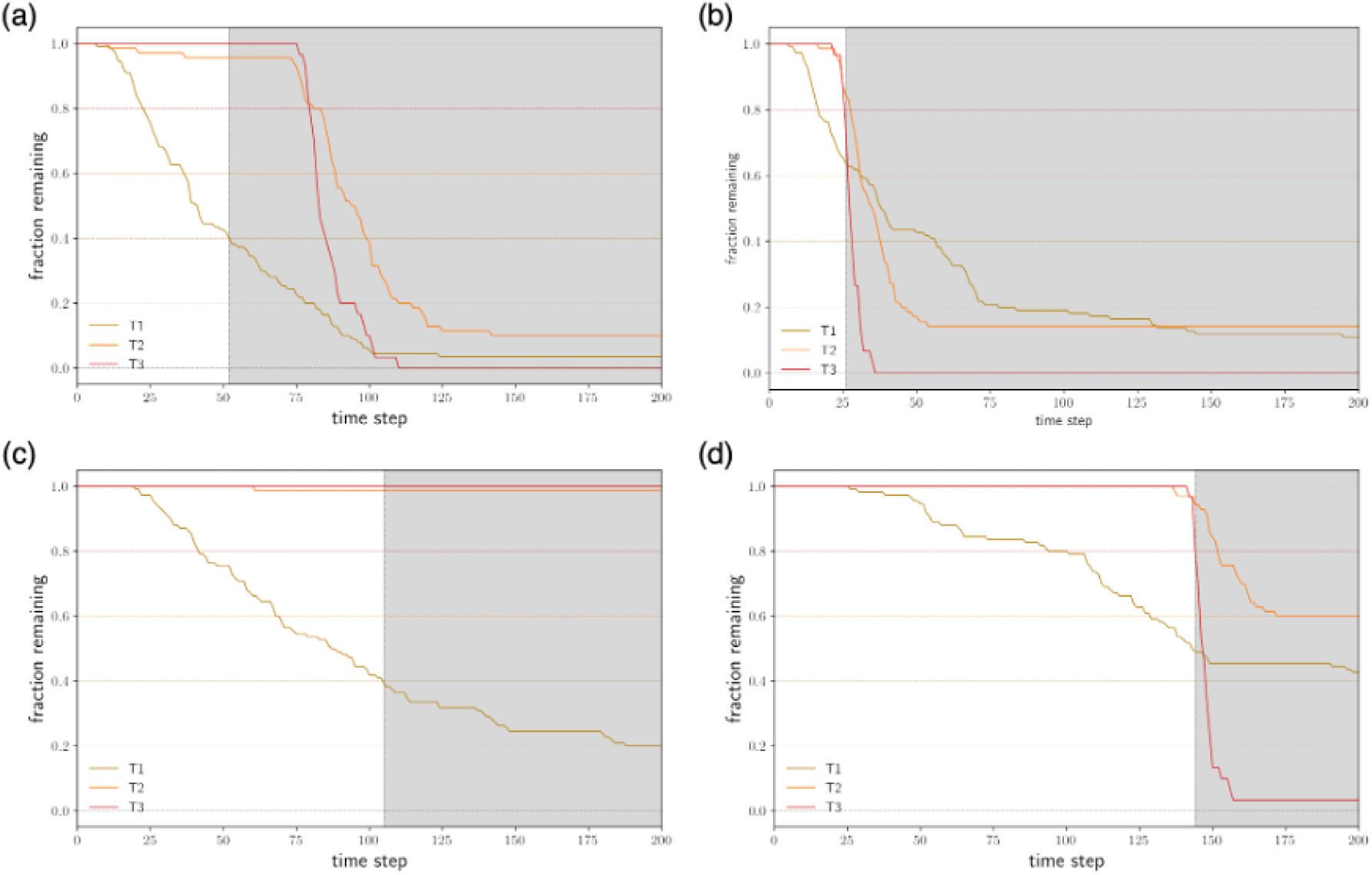
Time-to-implosion *T*_*imp*_ under increasingly protective regimes. Solid lines show tier-specific fractions of remaining members over time. Implosion is defined as the first time any of these fractions falls below its threshold (T1: 40%, T2: 60%, T3: 80%), denoted by the dashed lines. The non-shaded region marks the period that the bubble is viable; gray shading indicates time post-implosion. Bubble architecture, model processes, and model parameters are identical to those in Fig. 2 unless noted otherwise. In GP, recovery was disabled to maintain ongoing external transmission. (a) Weak protection: relational exchange rate *ζ* = 0.1; uniform boundary porosity *ω*_*i, j*_ = 0.6; no preferential mixing (*ϵ* = 0). (b) Moderately weak protection: as in (a) but with *ζ* = 0.3. Faster removal of infected members in this case actually leads to earlier implosion. (c) Moderately strong protection: as in (b) but with heterogeneous boundary porosities, identical to the bubble configuration in Fig. 2. (d) Strong protection: as in (c) but with preferential-contact matrix 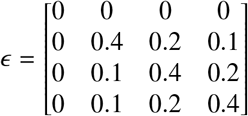, rows/columns ordered GP → T1 → T2 → T3.

This type of simulation-based analysis enables in-silico experimentation with bubble design and provides a robust framework for optimizing interventions across diverse outbreak scenarios to safeguard any predefined subpopulation.

## Discussion

The bubble framework intersects with foundational ideas in disease ecology and epidemiology on the epidemic effects of risk stratification. In classic studies of sexually transmitted infections, for example, hosts were stratified demographically by contact rate and susceptibility [Lajmanovich and Yorke, 1976], and “core” groups with disproportionate influence on transmission dynamics were subsequently identified for targeted interventions [Hethcote et al., 1982]. What distinguishes our framework is the deliberate manipulation of local population structures to achieve narrow public health aims, rather than modeling large-scale disease dynamics under naturally occurring demographic or behavioral variation.

The core principles of bubble defense are also widely evident in non-human systems. Many social animals notably employ spatial segregation as a pathogen-avoidance strategy; for example, wolves alter their infectious contact networks through group fission-fusion [Brandell et al., 2021], and female gorillas in polygynous groups have been documented leaving males with visible signs of illness to join younger, smaller groups [Baudouin et al., 2019]. Social insects display even closer analogues, protecting the colony from disease spread by modulating their social interaction networks, a process termed “organizational immunity” [Stroeymeyt et al., 2014], as part of a broader suite of defensive behaviors referred to as “social immunity” [Cremer, 2019]. As in human bubbles, division of labor gives rise to hierarchical organizations characterized by spatial compartmentalization and contact heterogeneity, which together creates a “fortress” colony that limits parasite access to prioritized members [Naug and Smith, 2007,Pie et al., 2004,Quevillon et al., 2015].

The bubble approach can be interpreted as a new variant of metapopulation theory, which, in disease contexts, predicts pathogen persistence in networks of geographically dispersed, semi-independent subpopulations in relation to local eliminations and host movement [Citron et al., 2021,Grenfell and Harwood, 1997,Keeling et al., 2004]. Although a bubble typically sits within a larger community, with members and non-members separated by short distances, its members-only regulations establish it as a semi-independent unit, creating a unique metapopulation-like system whose subpopulations remain in proximity.

Classical metapopulation models account for system heterogeneity in factors including patch quality, environmental disturbance, and local population density. Bubble models, by contrast, create heterogeneity to serve specific management objectives: individuals are sorted into priority-based tiers, permeabilities between tiers are adjusted hierarchically, contacts are assorted, and relational exchange is used to keep internal dynamics from mirroring those outside the bubble. Thus, by treating heterogeneity as design variables, the bubble approach brings strategic, goal-oriented architecture into traditional metapopulation thinking.

The bubble defense approach is particularly relevant in today’s fractured public-health landscape. During the multiple surges of COVID-19, vaccine hesitancy and inconsistent messaging from political leaders led to a lack of coordination and transparency, complicating mitigation efforts [Nagler et al., 2020,Sallam, 2021]. In certain countries, such structural problems have since become pronounced. Currently, in the United States, key public-health institutions are being hamstrung [Cutler and Glaeser, 2025], mistrust of scientific expertise is extensive [Tyson, 2024], and private funding for infectious-disease research has plummeted [Walrath, 2025]. Against this societal backdrop, the next epidemic could spread unchecked. However, bubbling can help defend essential or otherwise unprotected groups; if organized locally, it would function beyond the whims of political leadership, forming a set of local “firebreaks”. Our model framework offers strategic guidance and a rigorous foundation for planning, implementation, and evaluation. Its application is both scientifically and socially important: including bubble theory in scenario planning can enhance epidemic forecasts and help lessen communities’ entrenched reliance on top-down policies poorly aligned with local needs and constraints, replacing them with control measures that are strategically focused and autonomously governed.

### Methods (Bubble anatomy)

A basic representation of bubbles can thus be introduced by adding parallel epidemiological compartments – *S* _*B*_ and *I*_*B*_ – to the standard SIR (Susceptible-Infected-Recovered) framework. Such a model might assume that a) interactions among members are largely well-fixed; b) all members remain in the bubble indefinitely, except when expelled upon infection (at rate *ζ*); c) infections are detected and acted on promptly within the bubble: specifically, infected members are expelled before they can recover, hence no *R*_*B*_ compartment, and recovery (at rate *γ*) only happens to non-members; and d), for simplicity, the bubble population is replenished (at rate *ϕ*) by drawing from the general population’s uninfected pool, effectively the susceptible compartment for an emerging pathogen. These processes can be expressed as follows:

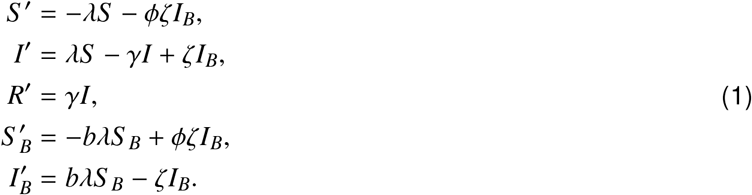

where *γ* is the recovery rate, and *λ* and *bλ* indicate the forces of infection outside and inside the bubble, respectively. The “strength” of a bubble is reflected in the relative force of infection, 1*/b*, and how quickly internal threats are removed, *ζ*.

### Boundary control

Bubble boundary restricts cross-tier mixing; we can introduce a porosity measure *ω*_*i, j*_ to denote the probability that an attempted contact between individuals from tiers *i* and *j* – one that would occur if no boundary existed – is realized. If the boundaries are completely impermeable but are active only periodically, *ω*_*i, j*_ can instead be read as the fraction of time the two tiers mix freely. In both cases, we can capture the result with a contact-structure matrix *κ* = [*κ*_*i, j*_], *κ*_*i, j*_ = *f* (*ω*_*i, j*_), where *f* is a monotonically increasing function. Each element *κ*_*i, j*_ is the share of a tier-*i* individual’s total attempted contacts across all tiers that ultimately connects with individuals from tier *j*; as boundary security tightens (*ω*_*i, j*_ decreases), *κ*_*i, j*_ falls.

We can define porosity as a constraint placed on cross-tier contact rates. The force of infection in tier *i* at time *t, λ*_*i*_(*t*), can then be given as

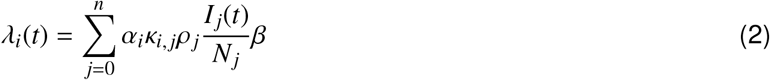

where *α*_*i*_ is the per-capita contact rate, *ρ*_*j*_ and 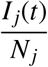 are the mean population density of tier *j* and the disease prevalence therein, respectively, and *β* is the probability of transmission upon infectious contact.

For simplicity, we may set *ω*_*i,i*_ = 1 and impose boundary symmetry *ω*_*i, j*_ = *ω*_*j,i*_. In a nested bubble, porosity between any two tiers would be the product of the porosities across each intermediate boundary:

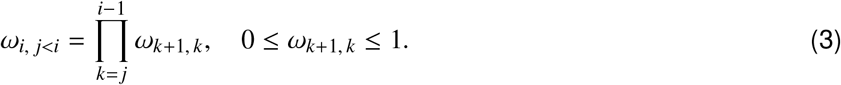

Since regulations could logically loosen near the bubble center (as in the NBA bubble), we may further assume *ω*_*k*+1,*k*_ ≥ *ω*_*k,k*−1_, *k* = 1, …, *n* − 1.

### Contact assortativity

We can adapt classic assortative-mixing models [Busenberg et al., 1990,Feng and Glasser, 2019,Glasser et al., 2012,Keeling et al., 2010] and express pairwise contact share *κ*_*i, j*_ as follows:

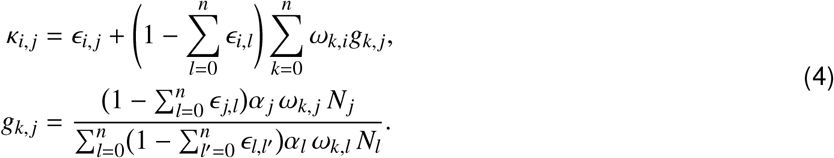

The first term *ϵ*_*i, j*_ ∈ [0, 1] represents preferential mixing: a fixed fraction of the total contact budget of a tier-*i* individual that is deliberately directed toward tier *j* partners, independent of contact opportunity. The second (summation) term captures proportionate (opportunity-driven) mixing; it distributes the remaining contacts (“floaters”) across all possible tiers *k* in proportion to member densities. Although Eq. (4) resembles earlier assortative models, it operates on the opposite premise: here, assortativity is not an emergent social pattern that management must accommodate, but rather an explicit tool for management. We can increase assortativity in a nested bubble by raising 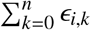 (thereby reducing random encounters) or reallocating preferential contacts so that only those that serve essential functions are preserved.

The preferential-contact matrix *ϵ* = [*ϵ*_*i, j*_] is comparable to the traditional WAIFW (Who-Acquires-InfectionFrom-Whom) matrix [Hilton and Keeling, 2019,Rohani et al., 2010,Schenzle, 1984]. However, instead of archiving observed rates of contact, *ϵ* specifies managerially imposed selective mixing that can be tuned *ex ante* to reshape transmission pathways. Because contact preferences need not be reciprocal, *ϵ* is generally asymmetric. Moreover, operational limit requires only that ∑_*j*_ *ϵ*_*i, j*_≤ 1 for every tier *i*; any residual share 1 −∑ _*j*_ *ϵ*_*i, j*_ remains available for proportionate mixing. By varying *ϵ* and modeling the epidemiological outcomes, scenario planners could then identify a distribution of preferential contacts that most effectively disrupts transmission chains linking the general population and high-priority members. For example, substituting random, unmonitored (proportionate) contacts with more frequent yet well-coordinated (preferential) contacts may significantly reduce the overall risk of pathogen importation (as would be reflected in *I*_*B*_).

### Imperfect compliance

Thus far, our analysis of bubble design has treated all members within a tier as behaviorally identical, each exhibiting the same level of responsibility and caution, an assumption unlikely to hold in practice (see Golliver [2021]). In general, nominally “equal-role” individuals differ in how strictly they adhere to bubble protocols, which can generate substantial within-tier variation in exposure risk and erode the protection afforded by secure boundaries and strategic assortment. Classical network theory (e.g., Dijkstra [1959], Newman [2001], Opsahl et al. [2010]) offers a natural way to address this heterogeneity in vigilance and its relationship to bubble effectiveness. Consider a set of bubble members *U*, which we represent as a weighted, undirected network. For any two directly connected members *x, y*∈ *U*, the edge weight (tie strength) *ψ*(*x, y*) is the risk of infection (or ease of transmission) associated with their contact; its inverse may be interpreted as a distance measure or resistance to transmission, e.g., disease spreads more slowly along longer or weaker ties. To incorporate individual “carefulness” into risk, we express *ψ*(*x, y*) as

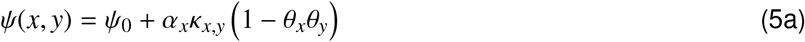

where *θ*_*x*_, *θ*_*y*_ ∈ [0, 1] are their respective adherences to presumably evidence-based health guidelines (0 = none, 1 = perfect), such as masking, handwashing, and social distancing, *α*_*x*_*κ*_*x,y*_ = *α*_*y*_*κ*_*y,x*_, and *ψ*_0_ *>* 0 is a non-zero floor of unavoidable risk between all dyad contacts.

Inside the bubble, let the shortest path (least-resistance) distance between nodes *u* and *u*^′^ be

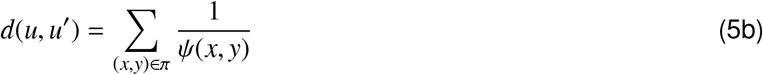

where *π*: *u* → *u*^′^ ranges over all ordered sequences of edges (*x, y*) linking successive members from *u* to *u*^′^. Let *U*_*b*_ ⊆ *U* denote those members who interact directly with the general population. For every node *u*, we can then define a risk-based centrality index

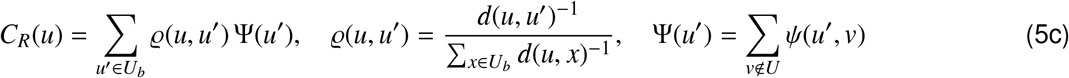

*C*_*R*_(*u*) combines conventional closeness centrality and degree centrality into a single risk score. Because a single rule-breaker can raise infection risk for everyone they can reach directly or through chains of contact, their presence tends to synchronize *C*_*R*_ among members who are spatially or socially proximate. In effect, heterogeneous adherence creates a risk gradient that stretches from the bubble perimeter to its interior. In a spatially explicit agent-based simulation model, the resulting *C*_*R*_ pattern could be captured by assigning each member a specific infection rate *β*_*u*_. A lapse in adherence at any node on the path linking two members both raises their individual *β* values and strengthens the correlation between them. Consequently, under realistic (imperfect) human behavior, clusters of similarly elevated *β* are expected to form throughout the bubble.

*C*_*R*_(*u*) can be mitigated by acting on its two key components: 1) pruning or removing high-risk edges between members and non-members reduces the perimeter contact term Ψ(*u*^′^), which operationally entails tighter boundary security (e.g., smaller *ω*_1,0_); 2) the weighting factor *ϱ*(*u, u*^′^) declines as the network distance between interior nodes and the perimeter *d*(*u, u*^′^) increases. In a nested bubble, stronger same-tier assortativity (e.g., larger *ϵ*_*n,n*_) lengthens these paths and thus dampens the impact of risky behaviors in other parts of the bubble.

Together, these measures can reshape the risk gradient of each tier, suppressing both pathogen importation and within-bubble spread.

Because *C*_*R*_(*u*) depends on membership, tier assignment, and evolving member behaviors (e.g., increasing laxity), the risk landscape is dynamic. At formation, identifying the member set *U* that minimizes long-term ∑ _*u*∈*U*_ *C*_*R*_(*u*) while still maintaining essential system functions yields an optimally composed bubble. Once formed, the bubble’s biosecurity can be refined through iterative edge reallocation, e.g., thinning optional contacts while reinforcing mission-critical ones. Periodic redirection of contacts may keep *C*_*R*_(*u*) low for high-priority individuals and shift residual risk toward peripheral zones where detection and intervention efforts can be concentrated.

## Acknowledgements

We thank Nathan Grubaugh and Shweta Bansal for their insights and contribution to our project discussion. We also credit the 2003-04 Detroit Pistons, whose toppling of the Los Angeles Lakers in the NBA Finals showed that success can come from relentless defense – victory through resilience.

## Author contribution

YT, CBO, and NB conceived and designed the research. YT developed the models and analyzed the results. All authors contributed to writing and editing.

## Data availability

Core Python code for generating the figures and simulation results is available at https://doi.org/10.5281/zenodo.17279695.

